# What Does It Take to Map a Country? Scaling OpenStreetMap Mapping for Accurate Health Accessibility Modelling in Madagascar

**DOI:** 10.64898/2026.03.25.26349339

**Authors:** Felana A. Ihantamalala, Masiarivony Ravaoarimanga, Mauricianot Randriahamihaja, Christophe Révillion, Lucas Longour, Tokiniaina Randrianjatovo, Feno H. Rafenoarimalala, Matthew H. Bonds, Karen E. Finnegan, Vincent Herbreteau, Fanjasoa Rakotomanana, Andres Garchitorena

**Author notes:** These authors contributed equally to this work (Felana A. Ihantamalala, Masiarivony Ravaoarimanga).

## Abstract

Comprehensive geographic data are essential to accurately model geographic accessibility to healthcare and to guide equitable health system planning and implementation. In low-income countries, however, incomplete road and building data in global databases such as OpenStreetMap (OSM) limit the precision and operational applications of geographic accessibility models. Following a successful pilot in one district of Madagascar, we evaluated the scalability of an exhaustive mapping approach to produce highly granulated household-level accessibility estimates at regional and national levels. Using satellite imagery and the OSM platform, we mapped all buildings, roads, footpaths, and rice fields across seven additional districts in southeastern Madagascar. We estimated travel routes, distance and travel time between each household and the nearest primary health center (PHC) or community health site (CHS) using the OSM Routing Machine, combined with predictions of travel speed from a locally calibrated statistical model. We then assessed population density and mapping completeness for roads and buildings in our study area and across Madagascar using AI-generated reference datasets (Microsoft and Facebook/MapWithAI) and estimated corresponding mapping times. Finally, we estimated the resources required in person-years to scale this approach across Madagascar using two different extrapolation methods. Nearly one and a half million buildings and 197,000 km of footpaths were added to OSM across the eight mapped districts, for a total area of about 30,200 km^2^. Between 24 % and 65 % of the population lived within one hour of a PHC depending on the district, and 87 %–99 % lived within one hour of a CHS. Most Malagasy districts were classified as having low completeness for both buildings and roads. Scaling up the approach to cover the entire country would require between 220 and 350 person-years depending on the extrapolation method and assumptions used. Mapping an entire country with sufficient detail to precisely model healthcare accessibility for every household is feasible but resource-intensive. Combining human mapping, participatory approaches, and AI-assisted datasets can substantially improve OSM completeness and generate actionable, high-resolution travel-time data for health planning. Our findings provide a roadmap for Madagascar and other countries seeking to develop national-scale geospatial infrastructure for sustainable development and universal health coverage.

## Introduction

Access to essential health services remains a significant challenge for nearly half of the world’s population [1]. In rural areas of sub-Saharan Africa, where health infrastructure is particularly sparse and the rates of primary health care access are among the lowest globally, geographic accessibility represents one of the main barriers to accessing care [2,3]. To address this, expansion of health infrastructure in underserved areas and last mile delivery interventions, such as mass distribution campaigns and community health programs, should ideally be optimized to the geographic challenges specific to the populations they target [4–8]. This operational optimization could be facilitated by integrating geospatial decision-making tools into the planning and implementation process.

Several methods exist to model geographic accessibility, ranging in complexity, precision, and data requirements. The simplest approach estimates Euclidean distances (i.e., “as the crow flies”) from health facilities using a constant travel speed, relying solely on facility location data [9]. More sophisticated methods integrate additional data, such as elevation, land cover, and road infrastructure, into least-cost-path algorithms, generating more precise estimates of travel times by accounting for variation in travel speeds across different terrains [3,9–12]. While these methods have proven useful in certain contexts, they often rely on generalized assumptions about travel routes and fail to provide precise, actionable information on the actual routes used by individuals. Routing algorithms can overcome these issues in settings with comprehensive geographic data on road networks and buildings, such as the Global North [13–16], but their application in rural areas of sub-Saharan Africa has been limited [17]. Of the many geographic databases of roads and buildings available, OSM has become one of the largest and most up-to-date open geographic databases at the global scale, comprising nearly half a billion buildings [18,19] and about 40 million kilometers of roads [20]. As a result, OSM data are widely used for navigation and mapping applications in multiple sectors such as disaster response, urban planning, healthcare and scientific research. OSM data are increasingly used in studies of geographic accessibility to healthcare in sub-Saharan Africa [17,21–24]. However, there remain wide disparities in the completeness of roads and buildings in the OSM database [23,28]. For instance, building completeness exceeded 70% in urban areas of Europe and Central Asia but it was only 30% in sub-Saharan African cities [25]. Similarly, road completeness is estimated to be nearly perfect in Europe and North America, but lower than 50% in many countries of sub-Saharan Africa and Asia [20]. Humanitarian mapping efforts have aimed to improve OSM completeness in low-income settings by adding millions of buildings and roads in the last decade, but these contributions still represent a small fraction of what is needed [18]. More recently, open AI-generated datasets developed by major technology companies such as Microsoft, Google, and Meta have attempted to close these data gaps. Yet comparative analyses in Africa and other low-income settings show large spatial disparities in completeness and accuracy, especially in rural areas, limiting their reliability for operational use in the Global South [19,26–29].

As an alternative, a pilot study combined a comprehensive participatory mapping of all roads, footpaths, and buildings on OSM in one health district of rural Madagascar with routing algorithms and locally-calibrated travel speed models to provide precise estimates of travel time to health facilities [17]. By estimating actual, detailed travel routes for both individuals accessing care and health workers accessing households, this approach allowed a move beyond simple time estimations, providing actionable information that can be integrated into e-health tools for operational decision-making. This not only enables real-time navigation but can also support health system and last-mile delivery planning in a variety of operational contexts [17]. This model has since been expanded for a variety of applications, including the accurate characterization of prehospital transport times [30], the optimization of community health post locations [31], and the planning and scheduling of last mile interventions that involve door-to-door delivery [32]. The aim of this study was to evaluate the scalability of this approach by applying it to seven districts in southeastern Madagascar, covering a population of over 2.5 million people and a total area approximately the size of Belgium. In addition to assessing the practical feasibility of expanding the approach regionally, we estimate the resources and time required to scale this approach across Madagascar, providing important insights into the viability of nationwide implementation in sub-Saharan African countries.

## Materials and methods

### Mapping and satellite data collection for health care accessibility models

To obtain the necessary data to model geographic barriers to care in our study area, a large-scale mapping project was carried out in three regions of southeastern Madagascar, in three distinct phases (Fig 1). The first phase was the above-mentioned pilot, conducted in Ifanadiana district from September 2019 to May 2020. This phase relied initially on volunteer contributions, but was primarily conducted (nearly 75%) by 5 members of the study’s team who worked part-time on it. The second phase was conducted in five districts of the Fitovinany and Atsimo Antsinanana regions: Manakara, Vohipeno, Farafangana, Vondrozo, and Vangaindrano. In this phase, ten full-time cartographers carried out the mapping from July 2022 to August 2023, and two coordinators supervised the work. The third phase mapped the remaining two districts of the Vatovavy region of which Ifanadiana is part: Mananjary and Nosy Varika. In this phase, four cartographers carried out the mapping from February 2024 to March 2025, and one coordinator supervised the work. Even though each of the mapping phases was part of a separate project with slightly different needs and purposes (see discussion), across the three phases, the team coordinating the mapping of the study area was the same, and the methodology used was consistent, as described below.

**Fig 1.**
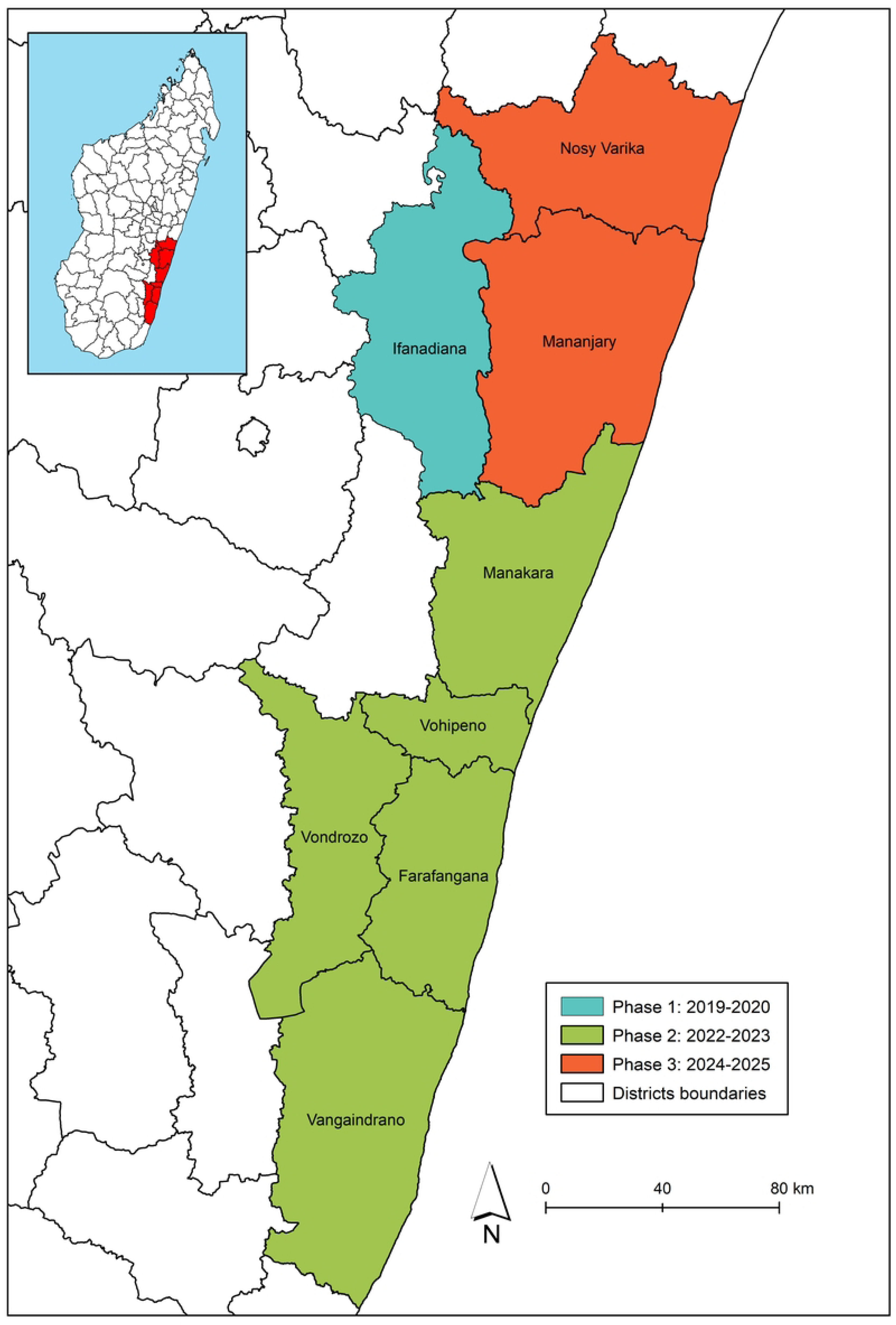
Geographic extent of the study area and timing of the OSM mapping, South-Eastern Madagascar 2019-2025. Each mapping phase is represented by a different color.

The mapping conducted in phase 1 in Ifanadiana district has already been documented [17], and the same methods were streamlined for scale-up in the seven additional districts during phases 2 and 3. Briefly, the study area was split into several mapping projects, each of which was in turn subdivided into tiles of 1km*1km. This resulted in about 30,000 individual tiles to be mapped separately (total study area was 30,200 km^2^) across the eight districts. For this, we used the Tasking Manager developed by the Humanitarian OpenStreetMap Team (HOT), a tool for coordination of volunteers and organization of groups to simultaneously and rigorously map on OpenStreetMap, especially during disaster response. Use of the Tasking Manager was initially done via a collaboration with HOT during phase 1, and then via a version of the Tasking Manager adapted by our team from the application’s source code published by HOT on GitHub for phases 2 and 3 [33] (S1 Table). This change was implemented because use of the Tasking Manager via HOT for large-scale projects was no longer free when phases 2 and 3 took place.

Mapping of each tile was done via photointerpretation of high-resolution satellite imagery available on OSM. For this, we used the ID editor directly on OSM during phase 1, and JOSM, an editor for OSM written in Java 11+, during phases 2 and 3 for more efficient edition (S2 Table). As background satellite imagery, we chose Bing for the phase 1 and phase 3 districts, and Maxar Premium for the Phase 2 districts. Mapping elements included all road networks of any size (including, but not limited to non-paved roads and footpaths largely absent from OSM), residential areas, buildings, rice fields and hydrographic networks (rivers, streams, water bodies). Mapping of each tile was done in two steps: first, one cartographer mapped the selected area, and then another cartographer went over the same area to look for missing geographical entities or to correct existing ones if necessary. This second step, validation, aims to provide OSM users with high-quality data. More details are available in Ihantamalala *et al.* (2020) [17].

In addition to generating and using OSM data, we downloaded satellite imagery and satellite-derived products to obtain land cover, precipitation and elevation in order to obtain context-specific estimates of travel time under dry and rainy conditions. For land cover classification, we obtained Sentinel-2 imagery from ESA’s Copernicus program with a spatial resolution of 10 m. Acquisition dates varied by district: August 18, 2018 for Ifanadiana; August 4, 2022 for Farafangana, Manakara, and Vohipeno; July 14, 2023 for Vangaindrano and Vondrozo. For Nosy Varika and Mananjary, we used multiple images to capture seasonal variation: dry season imagery from August 13, September 7, September 17, and October 2, 2023; and rainy season imagery from March 1, March 16, and April 25, 2023.

For the land use process, we used different software to perform a semi-automatic classification for phase 1, supervised classification for phase 2 and object-oriented analysis for phase3 (S3 Table). Initially, we manually delineated polygons representing regions of interest (ROIs) for each of the three land cover classes for phase 1 and 2: forest, water bodies, and savanna. For phase 3, we segmented the satellite images into meaningful objects and assigned the same land cover classes to each pixel with similar spectral value. Subsequently, we applied a different model that generates a response variable by constructing multiple decision trees and assigning each multi-layered pixel through every tree. We integrated two additional classes derived from OSM data: residential areas and rice fields. The land-use map was validated using field points collected and complemented by additional reference points identified through Google Earth for phase 1 and 2, and 20% of pixels used for the classification for phase 3 (S3 Table).

For precipitation, we obtained NASA Prediction of Worldwide Energy Resources (POWER), available at a spatial resolution of 0.5 x 0.625 degrees. For elevation, we obtained data from the Shuttle Radar Topography Mission (SRTM), downloaded from the United States Geological Survey (USGS), available at a resolution of 30 meters. From this elevation raster, we estimated a corresponding raster of slope using the slope function in ArcGis software.

Finally, a participatory mapping field campaign was conducted with the community health workers (CHWs) of three districts (Farafangana, Manakara and Vohipeno) to obtain the names of all hamlets and villages mapped on OSM during phase 2 and add them to the OSM database. Two field missions were conducted to Farafangana (March 22 to April 2 2022), and to Manakara and Vohipeno (July 16 to 31 2022). Participatory mapping took place during the monthly reviews of the CHWs at the PHC, facilitated by the head of the PHC and staff from local community health programs. With all the available OSM elements (buildings, rice fields, road networks, hydrographic networks, railroad lines, etc.), CHWs were given a paper map of their catchment area, which is defined as their respective fokontany (the smallest administrative unit and CHW catchment) to help them find landmarks and identify the villages/hamlets. Following a short explanation, CHWs were asked to write the names of all the villages/hamlets in their fokontany on the back of the map, number them, and then write each number in the corresponding location on the map.

### Estimation of distance and travel time to accessing health care

Once all buildings, roads and footpaths in the study area were fully mapped, we estimated the shortest path between each building in the study area and the nearest PHC or CHS. To do this, we used the OSM Routing Machine (OSRM) tool via the OSRM package in R software. OSRM uses Dijkstra’s routing algorithm, which iteratively searches for the shortest path between a unique node and the destination node in the network [34]. The departure point is each building in the OSM dataset, and the destination is the nearest PHC or CHS. The location and name of all PHC in the study area had been previously added to the OSM database. For CHS we assumed that they were located in the main village (“*chef-lieu*”) of each fokontany, as per Ministry of Public Health norms. For each household, both the shortest route and the corresponding distance were obtained and stored.

To estimate corresponding travel times for each of the shortest routes obtained through OSRM we applied variable travel speeds along the route based on local terrain characteristics. For this, each route was split into 100 m segments and each segment was intersected with land cover and slope layers. We then used the predictions of a statistical model of travel speed that uses these terrain characteristics as explanatory variables to obtain the total travel time of each route. This statistical model had been developed and calibrated during phase 1 with about 1000 km of GPS tracking fieldwork under diverse landscapes in Ifanadiana district [17]. Finally, to obtain a raster of distance and travel time, we performed kriging interpolation using ArcGIS software for the two types of health infrastructure.

To enable easy use and access of OSM data and geographic accessibility results by local health care actors, we created an online application for each of the three phases, as each phase was associated with a separate project and target actors. It consists on a web interface available in French and English developed with the Shiny package in R statistical software, and hosted on an internal Pivot server (research.pivot-dashboard.org for phase 1 and 3, mrp.pivot-dashboard.org for phase 2), which may be accessed for both private and public use (S1 Appendix).

### Data collection for estimation of National-Scale Mapping

To estimate the human resources required to achieve nationwide completeness of OSM data in Madagascar, we compiled and integrated several complementary datasets. These included available mapping statistics from our mapping efforts, existing OSM data, AI-generated reference databases, and population density layers.

First, for all the completed mapping projects in phase 2, detailed records from the OSM Tasking Manager were retrieved on the number of tiles mapped, together with mapping and validation times. These were used to calculate the average mapping time per square kilometer for each of the projects. In addition, an explicit mapping exercise was conducted to obtain precise tile-level mapping statistics on a sample of tiles in comparable settings. This mapping was carried out in four fokontany outside of the eight project districts selected for having the same level of completeness (see the next section below) as the phase 2 districts: Betsitindry (69.25 km²), in Port-Bergé district, Ankafanana (24.64 km²), Ambohitrambo (23.43 km²), and Ankazondandy (21.91 km²), located in the Arivonimamo district.

Second, building and road shapefiles available on OSM were obtained from the Geofabrik repository for three separate dates: January 2021 (before phase 2 mapping), September 2023 (after mapping of phase 2 was completed), and April 2024 (for the selection of the four fokontany). These were used to quantify completeness levels before mapping for phase 2 districts, and after mapping for the rest of Madagascar (excluding the study area).

AI-predicted features were incorporated to provide national-scale reference layers against which to assess completeness. These included Microsoft Building Footprints, which consist on polygon-level predictions of built structures obtained from the *GlobalMLBuildingFootprints* Github repository, and roads from MapWithAI/Facebook obtained from the *Open-Mapping-At-Facebook* Github repository, which consist on AI-detected transport networks. Third, gridded population estimates from *WorldPop* (2020) were used to estimate population density per district.

### Data analysis for estimation of National-Scale Mapping

To estimate the total resources required to map the entire country of Madagascar, two complementary methods were applied. First, mapping completeness was evaluated using Kontur’s *Disaster Ninja* framework, which compares OSM data to AI reference layers while accounting for population density [35]. Accordingly, each district was grouped into three levels of completeness: A represented low completeness (0 to 0.7), B represented medium completeness (0.7 to 0.9), and C represented high completeness (over 0.9). Each level of completeness was subdivided into three levels of population density: 1 represented districts with low population density (lower than 6.6 inhabitants per km^2^), 2 medium density (6.6 to 68 inhabitants per km^2^), and 3 high density (over 68 inhabitants per km^2^). As a result, each district was assigned to one of nine completeness subclasses (A1 to C3). However, while Kontur’s *Disaster Ninja* framework uses AI reference layers as the benchmark, in our setting these layers only represented a fraction of existing buildings (48%), roads and footpaths (8%). Thus, we estimated the ratio between the number of buildings and road length in AI reference layers versus the OSM database in our phase 2 districts (after mapping), which we used to adjust building and road completeness elsewhere in Madagascar. This classification was performed separately for buildings and roads at the fokontany level, providing a spatially explicit baseline for extrapolating mapping effort by district and completeness subclass. For each district, the proportion of land area in each completeness class was used to weight expected mapping effort. For each completeness class, median mapping and validation times per square kilometer were computed from Tasking Manager data for the classes observed in the mapped districts (A2, A3, B3) and then extrapolated to other classes using proportional relationships between successive classes (e.g., A1–A3, B1–B3, C1–C3). The total mapping time for each district was obtained as the sum of the area-weighted times across completeness classes. This method accounted for both baseline mapping completeness and population density, allowing estimation of the relative effort required across geographic contexts.

Second, using the tile-level dataset, a predictive statistical model based on mapping time per tile as recorded in the OSM Tasking Manager was carried out, where the sum of mapping and validation time for each tile (1 km²) was the response variable, and the explanatory variables were population density (inhabitants/km²), number of buildings per km², total road length per km², area of rice fields, and length of waterways. Tiles with missing or extreme values were excluded. For this, a multiple linear regression model was developed to predict mapping time (in seconds per km²) as a function of these variables. Variable selection was conducted through a stepwise procedure based on Akaike information criteria (AIC). The predictions from this model were then applied to all 113 administrative districts using national-scale layers for population density, road network, and buildings described above. Results were expressed in person-years, by assuming that one cartographer works 8 hours per day, 5 days per week, and 48 weeks per year.

## Results

### Large-scale OpenStreetMap mapping in south-eastern Madagascar

Overall, the three project phases resulted in a total of 1,145,500 buildings, 82,600 residential areas, over 176,000 km of footpaths and 168,000 ha of rice fields mapped (Table 1). The amount of mapping increased significantly from the pilot (phase 1) to the scale up phases (2 and 3). For instance, while 106,171 buildings were mapped in phase 1, 666,187 were mapped in phase 2 and 373,160 in phase 3. Similarly, 23,726 km of footpaths were mapped in phase 1, versus 97,560 in phase 2 and 55,047 in phase 3. Completeness in our study area before mapping campaigns started was generally low, with only 22.86% of buildings and 10.63% of footpath mapped. Substantial differences in mapping completeness were observed both between districts and between mapping elements. For instance, while over 50% of buildings had already been mapped in Vohipeno, less than 1% had been mapped in Vangaindrano. Similarly, completeness for secondary and tertiary roads was high for most districts, whereas footpaths were largely absent from the OSM database prior to our mapping campaigns, with less than 1% mapped in Ifanadiana, Farafangana, Vangaindrano, and Vondrozo. We observed similar heterogeneity for other land cover elements relevant to geographic accessibility modeling such as rice fields and waterways: completeness for rivers was substantially higher than for smaller streams, and completeness for rice fields prior to our mapping campaign was lower than 1% in Ifanadiana and Vangaindrano, whereas more than half had been mapped in Vohipeno and Nosy Varika. Following participatory mapping in collaboration with local CHWs in Manakara, Vohipeno and Farafangana districts, a total of 4870 village names were added to the OSM database, which represented over three quarters of the currently available village names in these districts.

**Table 1:**
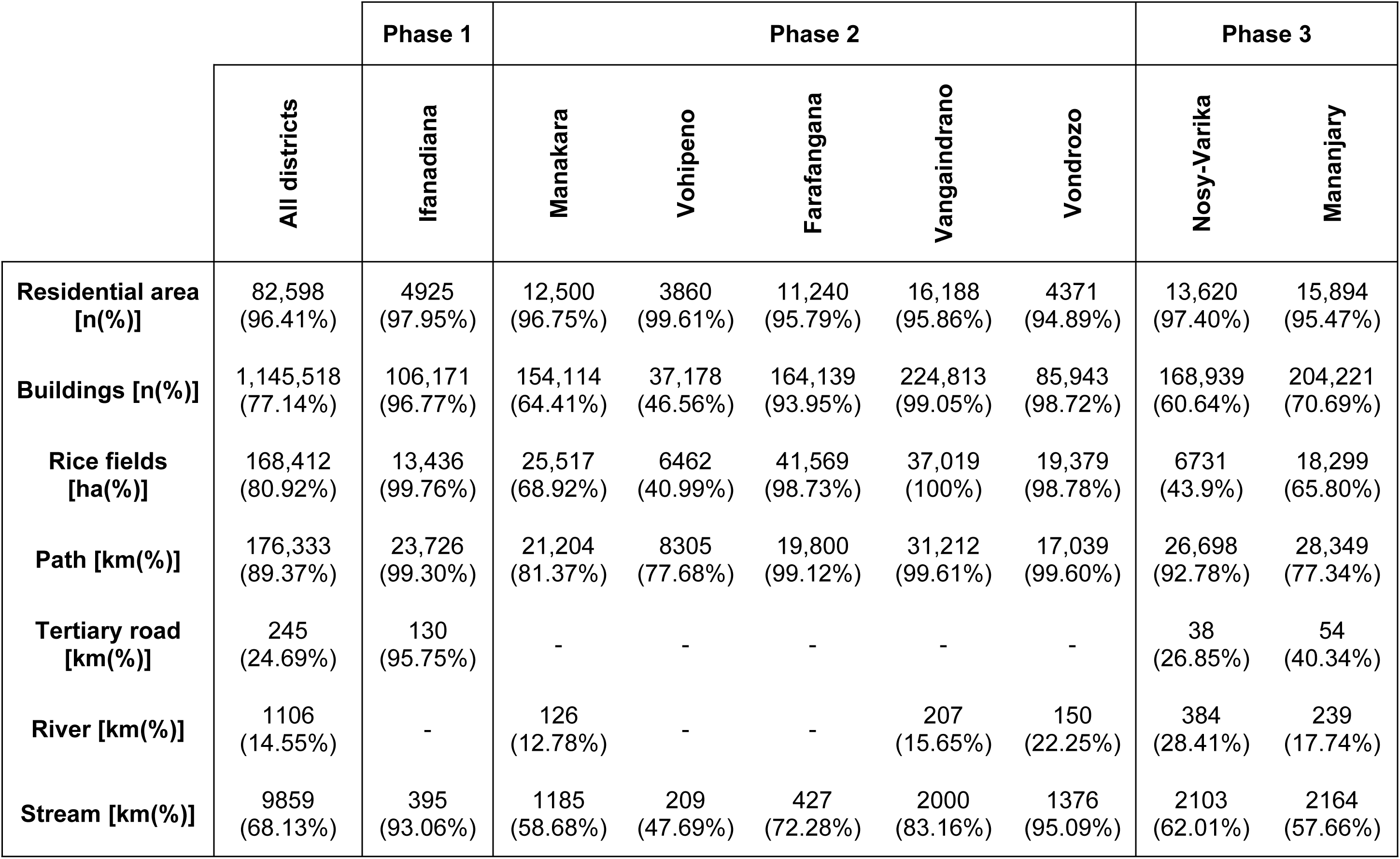
Data added to the *OpenStreetMap* database during pilot (phase 1) and scale-up (phases 2 and 3) mapping campaigns. Results are given in terms of elements mapped in our study (number, surface or length) and the percentage of the total available (mapped in our study).

### Travel time to reach the nearest community health site and primary health care center

Locally-calibrated geographic accessibility estimates for the nearly 1.5 million buildings in our study area revealed that geographic access to primary care facilities remains low in south eastern Madagascar. Overall, less than 42% of the population in the study area lives within one hour from a PHC in the dry season and less than 40% in the rainy season (S1 Table). Over 60% of the population in Ifanadiana, Vondrozo, Nosy-Varika, and Mananjary live more than 1 hour from the nearest PHC, and between 20 and 35% live more than 2h away (Table 2 and Fig 2a). In contrast, access to CHS is considerably better, with more than 90% of the population across all districts living within 60 minutes of a CHS in both dry and rainy seasons. Geographic accessibility to CHSs is particularly high in Farafangana, Manakara, and Vohipeno, where over two-thirds of the population live within 30 minutes of a CHS (Fig 2b). However, accessibility challenges remain at the community health level for Ifanadiana, Mananjary, Vangaindrano and Vondrozo, where about 10% of the population lives more than 1h from the nearest CHS with or without rainfall.

**Fig 2.**
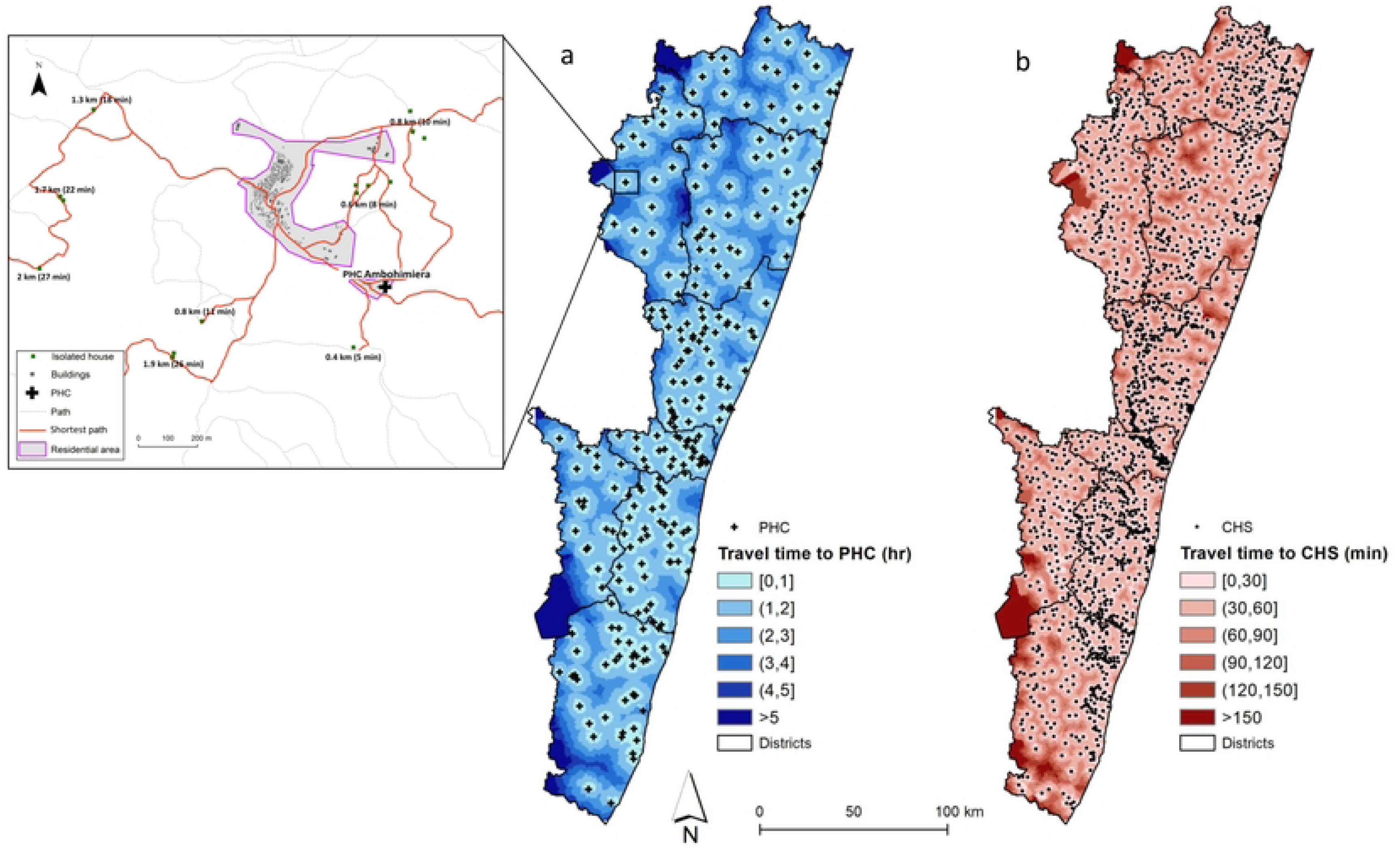
Spatial distribution of geographic accessibility to primary care in south-eastern Madagascar. Maps show estimations of travel time without rainfall to the nearest (A) primary health care center (B) community health site for the nearly 1.5 million buildings in the OSM database.

**Table 2:**
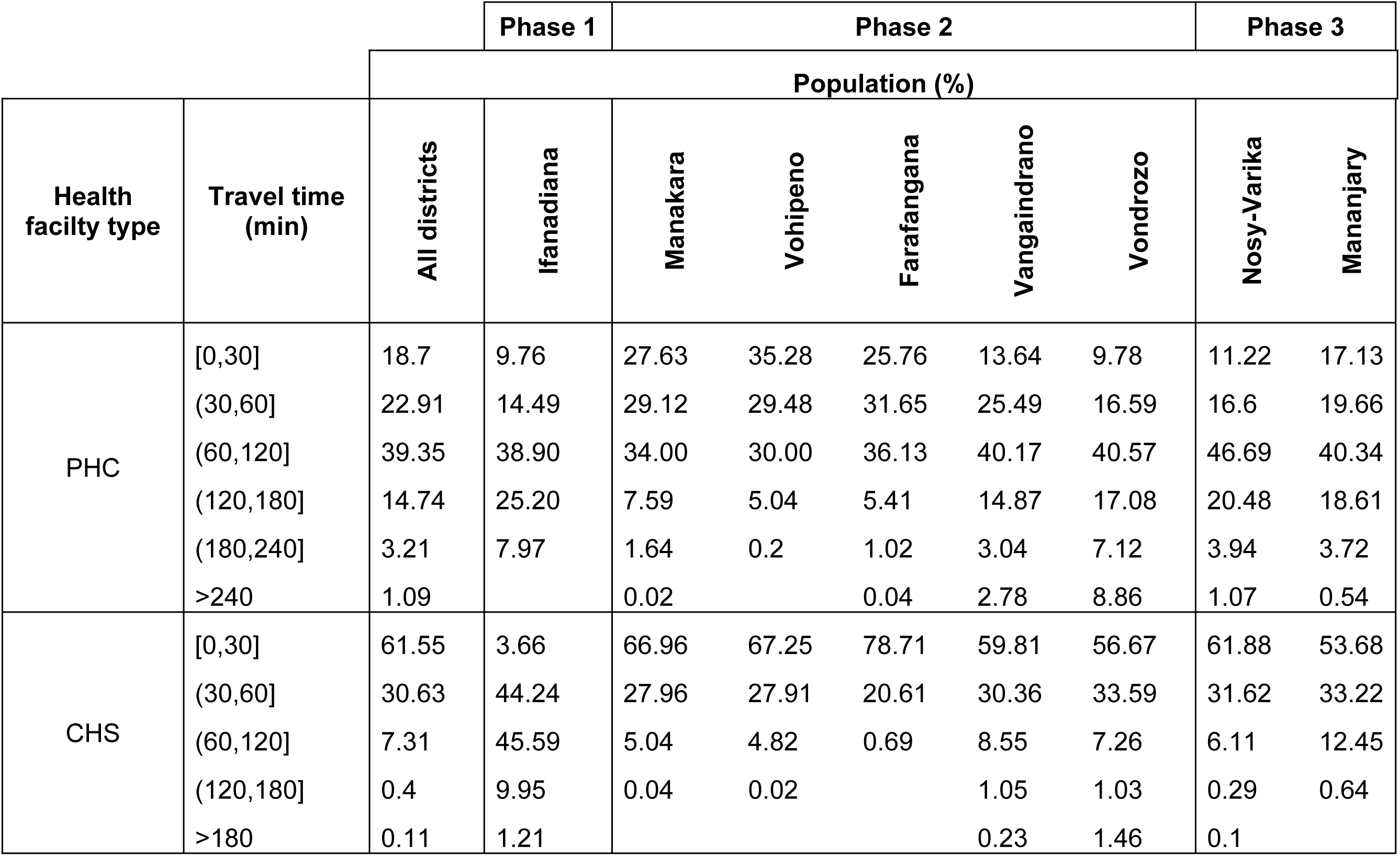
Distribution of the population (%) by travel time without rainfall to the nearest health facility. Results are disaggregated by access to Primary Health Centers and Community Health Sites, across eight districts in southeastern Madagascar.

|  |  |  | Phase 1 | Phase 2 |  |  |  |  | Phase 3 |  |
| --- | --- | --- | --- | --- | --- | --- | --- | --- | --- | --- |
|  |  | Population (%) |  |  |  |  |  |  |  |  |
| Health facility type | Travel time (min) | All districts | Ifanadiana | Manakara | Vohipeno | Farafangana | Vangaindrano | Vondrozo | Nosy-Varika | Mananjary |
| PHC | [0,30] | 18.7 | 9.76 | 27.63 | 35.28 | 25.76 | 13.64 | 9.78 | 11.22 | 17.13 |
|  | (30,60] | 22.91 | 14.49 | 29.12 | 29.48 | 31.65 | 25.49 | 16.59 | 16.6 | 19.66 |
|  | (60,120] | 39.35 | 38.90 | 34.00 | 30.00 | 36.13 | 40.17 | 40.57 | 46.69 | 40.34 |
|  | (120,180] | 14.74 | 25.20 | 7.59 | 5.04 | 5.41 | 14.87 | 17.08 | 20.48 | 18.61 |
|  | (180,240] | 3.21 | 7.97 | 1.64 | 0.2 | 1.02 | 3.04 | 7.12 | 3.94 | 3.72 |
|  | >240 | 1.09 |  | 0.02 |  | 0.04 | 2.78 | 8.86 | 1.07 | 0.54 |
| CHS | [0,30] | 61.55 | 3.66 | 66.96 | 67.25 | 78.71 | 59.81 | 56.67 | 61.88 | 53.68 |
|  | (30,60] | 30.63 | 44.24 | 27.96 | 27.91 | 20.61 | 30.36 | 33.59 | 31.62 | 33.22 |
|  | (60,120] | 7.31 | 45.59 | 5.04 | 4.82 | 0.69 | 8.55 | 7.26 | 6.11 | 12.45 |
|  | (120,180] | 0.4 | 9.95 | 0.04 | 0.02 |  | 1.05 | 1.03 | 0.29 | 0.64 |
|  | >180 | 0.11 | 1.21 |  |  |  | 0.23 | 1.46 | 0.1 |  |

### Estimated OpenStreetMap completeness in Madagascar for roads and buildings

Assessment of OSM data completeness, revealed very low levels of completeness both for roads and buildings in Madagascar (Table 3). Over 90% of districts were classified in the A category (completeness under 0.7), with the most prevalent class being A2 (medium population density) for both road completeness (51% of districts) (Fig 3a) and building completeness (58% of districts) (Fig 3b). Furthermore, 34% of districts fell into class A3 for building completeness and 13% for roads, corresponding to areas with high population density but very low OSM data completeness. Only 4 districts were classified as having high building completeness and 15 as having high road completeness (class C, over 0.9).

**Fig 3.**
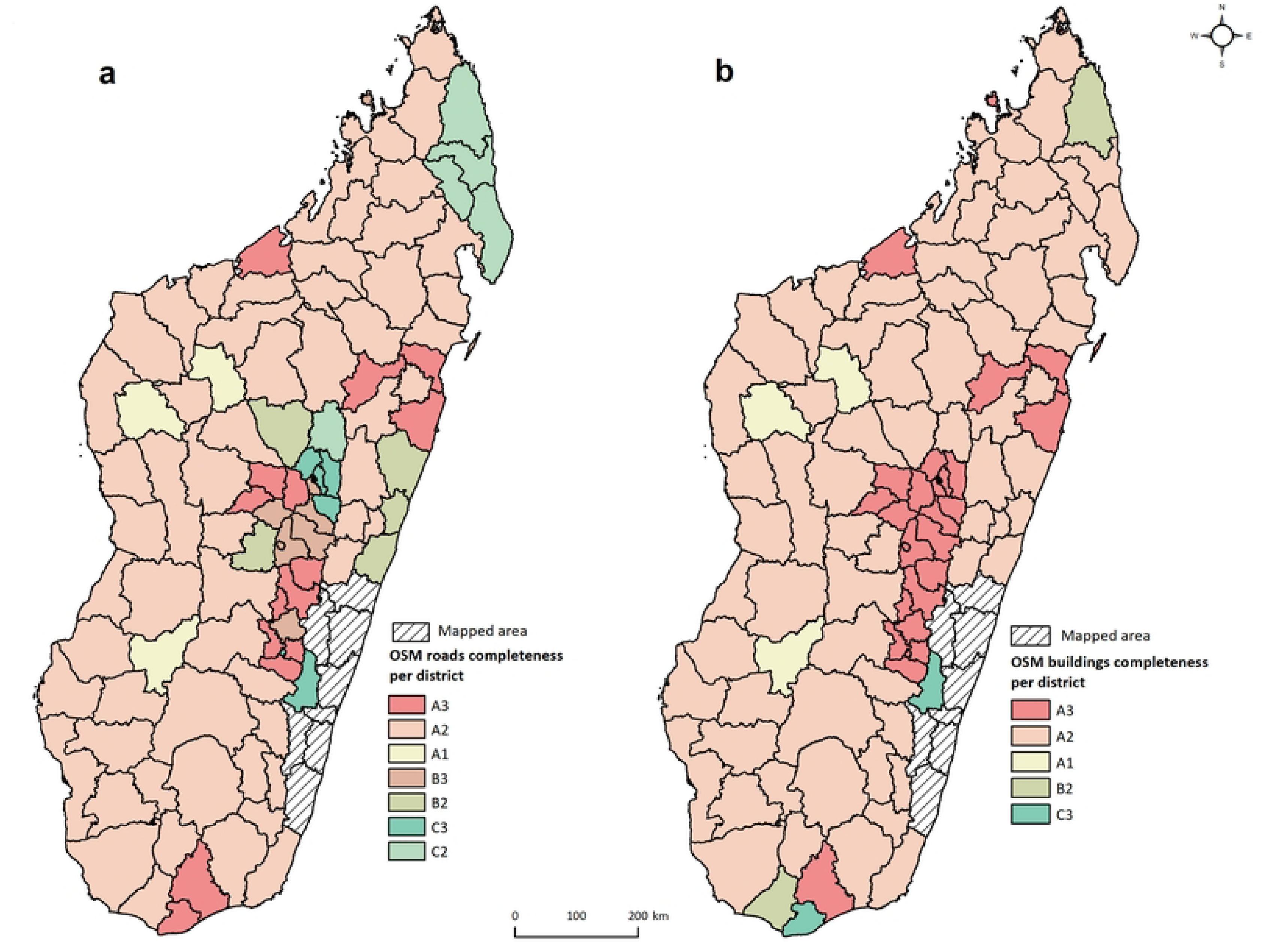
OpenStreetMap completeness per district for roads and buildings in Madagascar. (A) Map on left shows completeness classes for roads. (B) Map on right for buildings, based on Kontur’s Disaster Ninja framework.

**Table 3:**
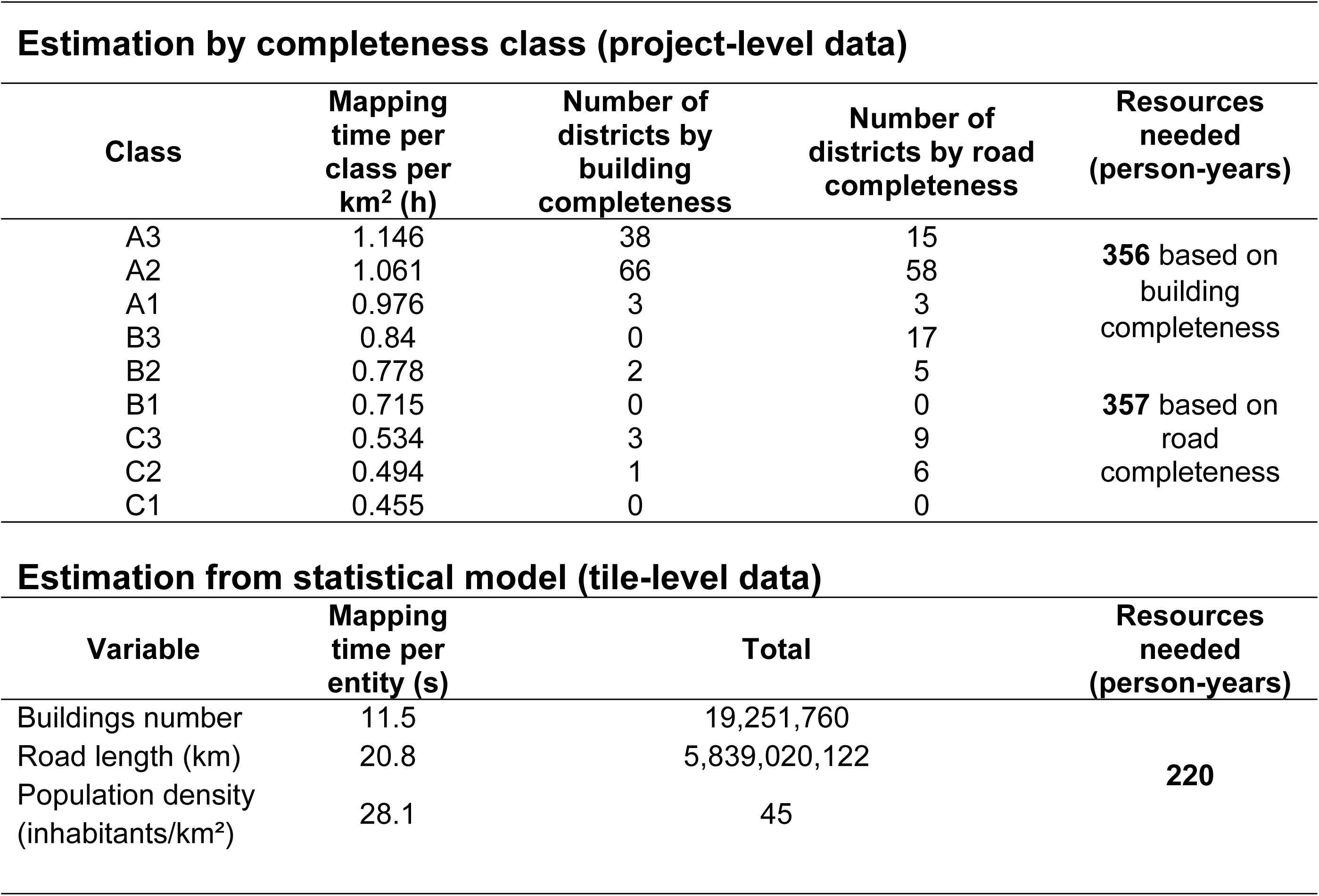
Estimation of the resources needed to map Madagascar based on analyses of project-level and tile-level data

### Resources needed to map Madagascar on OpenStreetMap

Analyses of mapping time (mapping + validation) per completeness class and square kilometer during phase 2 revealed that mapping time varied widely according to density and completeness (Table 3). For instance, while it only took about 30 minutes to map 1 km^2^ in areas of high completeness (C class), it took 1 hour to map areas of low completeness (class A). Given the same level of completeness, high density areas took about 20% longer to map than low density areas. Based on these results together with districts’ surface area and corresponding class, we estimated that it would require about 350 person-years to map all of Madagascar whether we used building or road completeness data (Table 3).

Results from the statistical model of tile-level mapping time, which included the use of AI-generated datasets in the mapping routine, revealed that three variables had a statistically significant association with mapping time: the number of buildings, the road/footpath length and population density of the tile (Table 3). Based on this model, we predicted that mapping a 1 km^2^ tile takes 11.5 seconds for every additional building, 20.8 seconds for every 100 meters of additional road/footpath, and 28.1 seconds for an increase in population density of 10 inhabitants per km^2^. Extrapolating these results by district and across Madagascar, we estimated that it would take 220 person-years to map all of Madagascar. Predicted mapping time was highly heterogeneous across the country. About one quarter of districts showed high predicted mapping times, in excess of 4500 hours. This was especially the case for those located in the south, part of the highlands (center) and north-west of the island (Fig 4). Conversely, several districts along the east coast had estimated times under 1000 hours. Intermediate districts with predicted mapping time between 1000 and 4500 hours were mostly located in the transition zones between major urban centers and less populated rural areas.

**Fig 4.**
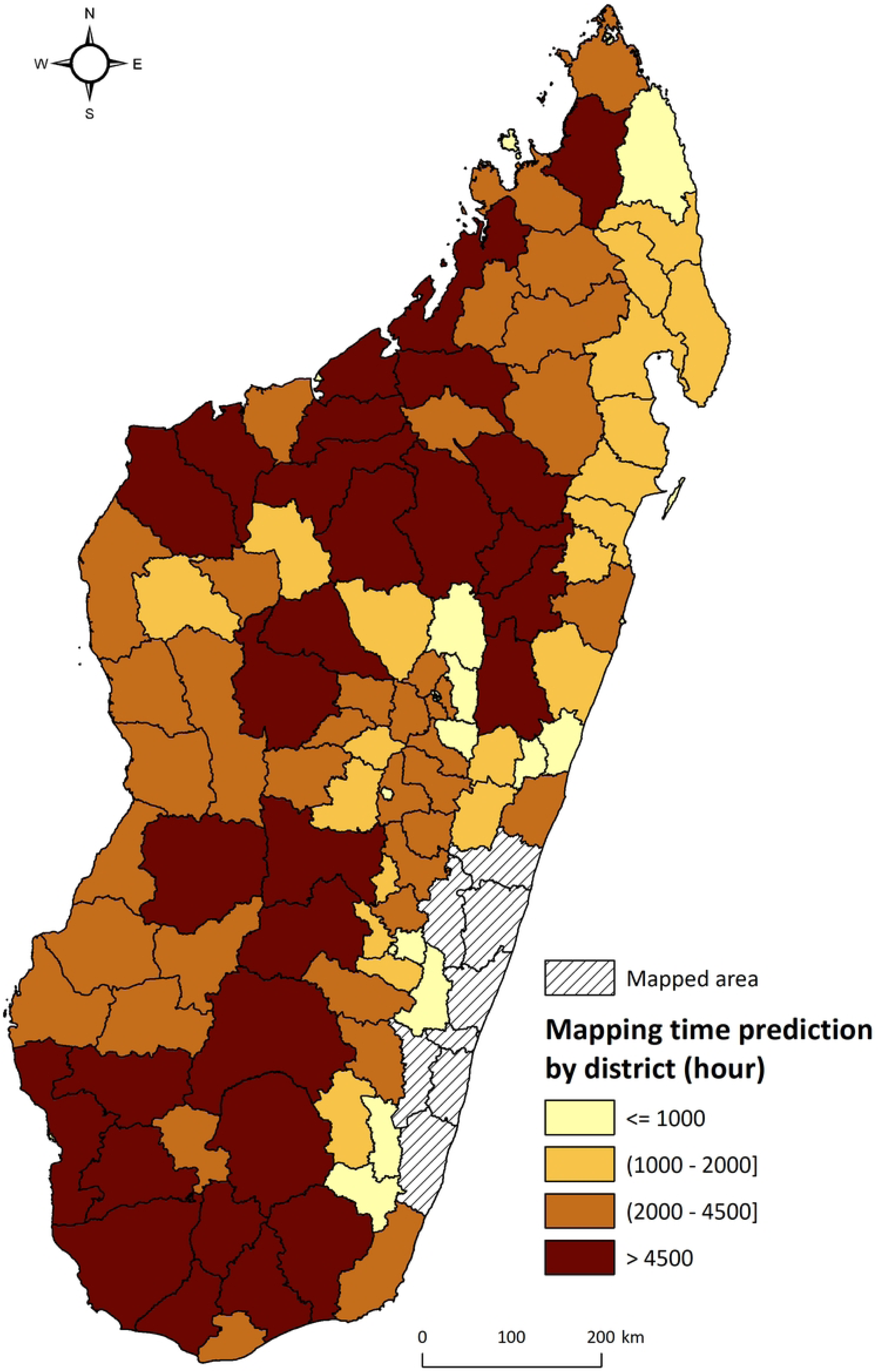
Prediction of mapping time by district in Madagascar. Colors represent the time needed for complete mapping, classified into four classes: ≤ 1000, 1000-2000, 2000-4500 and >4500 hours. Districts in white represent districts already mapped in this study.

## Discussion

*OpenStreetMap* has become a reference dataset for geospatial analyses, with operational applications across a wide range of fields, thanks to the richness and completeness of its data, open access and simplicity of data reproduction [35,36]. In health care, OSM data is increasingly used in geographic accessibility modeling, and it has allowed for applications such as routing and scheduling of home care delivery in the US and Europe [3,37]. However, low mapping completeness in rural areas of the Global South prevents integration of these data into operational applications to improve public health programs [22,28,34]. In this study, we scaled up a pilot program that combines complete OSM mapping of entire districts with locally calibrated estimates of travel time to build operational decision-support tools for community health programs. By mapping 30,200 km^2^ across 8 districts of southeastern Madagascar between 2019 and 2025, we demonstrate the feasibility of implementing this approach at scale and estimate the resources needed to scale it across Madagascar. Using this approach, we show that 58.39% of the population in the study area are located further than 1h from a PHC and 7.82% are located further than 1h from a CHS. By extrapolating information from our regional scale-up to all of Madagascar, where data completeness is low across the country, we estimated that between 220 and 357 person-years would be necessary to map the island.

By providing detailed and freely accessible spatial information on infrastructure, transportation networks, land use, and public amenities, OSM is a valuable resource for spatial analyses and planning across sectors [38]. In areas where completeness is high, OSM can be used in public health initiatives, providing essential geospatial data for disease surveillance, assessments of healthcare access, and coordination of emergency response operations [39]. In addition, OSM has proven valuable in environmental health research, enabling the identification of pollution sources and the assessment of green space distribution, helping to promote healthier and more resilient communities [40]. Applications from other sectors could also serve as inspiration for health care initiatives in the global south. For instance, urban planners use OSM data to optimize transport infrastructures and improve equitable access to essential public services [41,42]. Furthermore, OSM use is widespread in service and product delivery applications, from garbage collection to package delivery [37]. Our approach applies these methods to inform health programs in rural Madagascar, by estimating travel routes for households and health workers in fully mapped areas to provide actionable information for public health decision-making. This includes characterization of access to community health, health centers and hospitals based on household-level information to optimize the construction of new infrastructure [17,30,31], together with real-time navigation during field missions and the optimization of door-to-door delivery interventions to support community health programs [17,32]. However, the real-world use of this approach at scale across sub-Saharan Africa is highly dependent on the level of OSM completeness and on the feasibility of improving completeness over relatively short time frames.

We show that over 90% of Madagascar remains poorly mapped, and that mapping time varied substantially depending on road and building completeness, and population density. Recent research on OSM completeness highlights both the lower OSM completeness in sub-Saharan Africa as compared to other areas, and the methodological challenges for accurately assessing completeness. It is estimated that many sub-Saharan African countries have road and building completeness below 50% as compared with near complete mapping of areas in Europe or the US [20,25]. However, different completeness measures can yield substantially different results, with count ratios tending to underestimate building completeness while area ratios tend to overestimate it [43]. To address the challenge of estimating OSM completeness without reference data, researchers have developed proxy indicators [44,45]. For instance, building density has shown an approximately linear relationship with building completeness [44], while geometric indicators like area, perimeter, and density can accurately estimate street block completeness [45]. In our study, we relied on a commonly used method in disaster response mapping, which consists of classifying areas according to both their population density and their level of completeness using available AI-generated datasets as reference [35]. We found that completeness across most of Madagascar was very low for both buildings and roads, although road completeness was higher than building completeness. Correspondingly, estimated mapping time for a country as large as Madagascar (587,041 km², larger than metropolitan France for example) was high: we predict that it would take 100 full-time mappers 2 to 3.5 years to complete the mapping.

The development of AI and its use in the creation of OSM data, which is making mapping by human contributors faster and more accessible, holds promise for substantial and rapid improvements in OSM completeness [46,47]. Indeed, AI can automatically identify elements such as buildings, roads, and rivers in satellite imagery, enabling cartographers to cover vast areas that were previously poorly mapped or unmapped in a short time [48]. Currently, however, AI still faces important limitations that prevent it from fully replacing human contributors. AI models often misinterpret satellite imagery, confusing shadows, trees, or temporary structures for buildings or roads, and they struggle with smaller or irregular features such as footpaths or informal settlements, widespread in rural areas of the global south [49–51].

AI performance is closely dependent on the quality and release date of training and prediction data, such as available satellite images and corresponding elements mapped on the OSM database by human contributors. Thus, AI-generated data needs to be carefully validated, as large-scale automated modification could compromise the reliability of data on OSM. Recent studies in Africa and other low-income settings revealed substantial inconsistencies across rural and urban contexts between AI-generated datasets developed by major technology companies, introducing major uncertainty for programs aiming to use them [26–29]. For instance, the choice of AI-generated dataset for the identification of priority settlements for indoor residual spraying (IRS) campaigns in Zambia resulted in a 230 % variation in the number of identified candidates [28]. In our study area, MapWithAI/Facebook road data only identified a small fraction of footpaths, while use of Microsoft building footprints revealed frequent discrepancies when compared with available satellite images on OSM. As a result, to use these AI-generated datasets as reference data for completeness estimations, we had to scale MapWithAI/Facebook road length predictions by a factor of 11.67 and Microsoft building count predictions by a factor of 2.05. Moreover, the use of these data in the tile-level mapping exercise resulted in a lower prediction of total resources (220 vs. 357 person-years), but it is unclear whether this was due to the use of AI-datasets or other factors (estimation method, individual differences in mapping speed, etc.). In practice, we found that use of AI-generated data for mapping it was only moderately helpful in our setting.

As an alternative to unreliable AI-generated datasets and insufficient contributions from volunteers in areas of the Global South not affected by natural disasters or other emergencies, we hired full-time professionals to carry out the mapping, while taking advantage of existing collaborative platforms to parallelize and coordinate their work. A simple extrapolation of person-years into costs based on a typical salary for Madagascar ($300 per month) suggests that mapping Madagascar in such a way would cost about one million US dollars (+/-20%), with additional costs for coordination, supervision and materials. These costs should be put in perspective with the potential benefits that a fully mapped country and related geospatial applications could have, not only for the Ministry of Public Health and humanitarian organizations, but also across multiple sectors related to human development (urban planning, education, etc.). There are many potential applications, especially because OSM data is freely accessible and can therefore be used by all citizens. Moreover, by substantially increasing the amount of quality, human-validated data in a country from the global South, this could result in an increased ability by AI-models to predict OSM features in these settings, further helping to close the gap in OSM completeness elsewhere.

Our study had some limitations. First, the estimation of travel time across the eight districts considered variations in travel speed according to rainy versus dry conditions, but did not consider broader accessibility challenges due to flooding. Moreover, travel speed was locally calibrated from data recorded by healthy individuals, and did not take into account variations in travel speed according to patient profiles such as sick individuals, the elderly, or pregnant women. Second, estimates of geographic accessibility are based on individual itineraries for about 1.5 million buildings, but satellite imagery does not distinguish between inhabited households among buildings, which can lead to inaccuracies in population-level estimates. Finally, we could not obtain data on mapping time for individual tiles from the Tasking Manager during our mapping campaigns, as this information was only available at the level of the task/project. As a result, estimation of national-scale resources used two separate methods and data sources: aggregated project-level data of the whole area mapped during phase 2, and disaggregated tile-level data from a dedicated exercise in a much smaller area of similar characteristics, which yielded very different estimates (350 vs 220 person-years). This could be partly due to the limited variation in completeness classes in phase 2 mapping, which resulted in extrapolation of mapping time for other classes that may not accurately reflect true values. Moreover, the A class in Kontur’s groups together areas with very different levels of completeness (0-0.7), which may have resulted in an overestimation of mapping time for districts that were on the higher end of the completeness range as compared to phase 2 districts. Furthermore, we used the completeness of AI-generated datasets in our phase 2 districts to obtain district completeness classes elsewhere, which may have also led to an overestimation of mapping time if these datasets have higher completeness in other areas of Madagascar. Analyses based on a larger sample of settings with different characteristics (urban, peri-urban, rural, desert area with different types of landscape and levels of completeness) could help improve the accuracy of mapping time predictions in future studies.

## Conclusion

Achieving equitable access to healthcare requires first knowing where people live, how they move, what barriers they face, and how to get to them. Our work illustrates that building this spatial knowledge at scale on open databases remains a major but achievable challenge for countries of the Global South. By coupling exhaustive *OpenStreetMap* mapping with AI-generated reference datasets and predictive models of mapping effort, we provide a reproducible framework for estimating the human and temporal resources required to map very large areas in data-scarce settings. Such an approach could enable informed investments in geospatial infrastructure and contribute to a broader research agenda at the intersection of data science, participatory mapping, and global health. Together, improvements in OSM completeness at the national or regional level could enable ministries, humanitarian partners, and researchers to integrate high-resolution accessibility models into operational planning and implementation—accelerating the way toward data-driven decision-making for universal health coverage and beyond.

## Supporting information

**S1 Table:** Different Tasking Manager used for the three projects

**S2 Table:** Comparative visualization interface for OpenStreetMap

**S3 Table:** Methods used in the remote sensing process for the three projects

**S4 Table:** Distribution of the population (%) by travel time with rainfall to the nearest health facility. Results are disaggregated by access to Primary Health Centers and Community Health Sites, across eight districts in southeastern Madagascar.

**S1 Appendix:** Methodology for estimating distance and travel time to access healthcare

## Data Availability

Data are available on OpenStreetMap (https://www.openstreetmap.org) and on the Shinny app described in both studies https://research.pivot-dashboard.org/ and https://mrp.pivot-dashboard.org/

https://research.pivot-dashboard.org/

https://www.openstreetmap.org

## Acknowledgements

We gratefully acknowledge the support of USAID Access in Manakara and Vohipeno. We thank NGO Inter Aide Farafangana, especially Mrs Nadia and her team, for their valuable collaboration. Our appreciation extends to the chiefs of the Primary Health Centers in Manakara, Vohipeno, and Farafangana, as well as the Community Health Workers, for their essential collaboration. We also recognize the dedicated work of all mappers who participated in the three phases of the project.

